# Climate Care India: A protocol for digital transformation of health systems for non-communicable disease management and climate change adaptation

**DOI:** 10.1101/2025.05.25.25328322

**Authors:** Jasmin Bhawra, Sheriff Tolulope Ibrahim, Jamin Patel, Anuradha Vaman Khadilkar, Tarun Reddy Katapally

**Author notes:** **Corresponding author:** Tarun Reddy Katapally.

## Abstract

**Background and objective:** Non-communicable diseases (NCDs) account for 74% of global deaths, disproportionately affecting low-resource settings in the global south. The increasing frequency and severity of climate change-related events worsen the NCD burden – particularly in low-resource settings – thereby necessitating health system transformation. This longitudinal trial aims to transform the current response to climate change and NCDs among affected communities via a customized digital health platform.

**Methodology:** Building on the intergenerational Youth Adolescents’ behaViour, musculoskeletAl heAlth, Growth & Nutrition (YUVAAN) prospective cohort study – which has enrolled 1070 rural households as of November 2024 in Western India – a digital platform will be tailored to monitor and address evolving climate change and NCD-related risks. The three-phase methodology includes: 1) adaptation: co-developing the platform with a Citizen Scientist Advisory Council for local use, integrating culturally relevant features and languages; 2) implementation: pilot testing the platform and step-wedge deployment within the YUVAAN cohort; 3) evaluation: conducting mixed-method analyses of the platform, climate change, and health outcome associations. A sample size of 978 families was calculated to detect an effect size of 0.3 (90% power, 0.05 α, 15% attrition).

**Discussion:** Through access to real-time data, the digital platform will provide rural households with personalized support for NCD prevention and management, while enabling climate change preparedness and adaptation strategies in participating communities.

**Conclusion:** Integrating digital platforms into local decision-making will strengthen health systems’ capacity to manage NCDs in rural and low-resource settings impacted by climate change. These platforms can enable real-time data access for personalized care and evidence-based decision-making.

## 1. Background

Non-communicable diseases (NCDs), including diabetes mellitus, cancers, cardiovascular diseases, and chronic respiratory diseases, are a leading cause of all deaths globally [1]. NCDs arise from a combination of risk factors, including behavioural [2], metabolic [3], and environmental factors [4]. Behavioural risk factors include modifiable lifestyle choices such as tobacco use, unhealthy diet, physical inactivity, and harmful alcohol consumption that contribute to NCDs [5]. Metabolic risk factors, including physiological conditions such as high blood pressure, elevated blood glucose, abnormal lipid levels, and obesity, are also major contributors to the development and progression of NCDs [6]. Environmental factors, including climate change-related events such as rising temperatures [7], extreme weather conditions [8], and worsening air pollution [9], significantly increase the risk of respiratory diseases, cancers, and cardiovascular conditions, particularly in regions with inadequate environmental regulation and protections and an increased population [10]. These risk factors are deeply interconnected [11]; thus, they must be addressed concurrently to reduce the overall burden of NCDs [12].

Among the various NCDs, cardiovascular disease, asthma, and respiratory disorders are associated with higher mortality rates [13]. Cardiovascular diseases alone account for approximately 17.9 million deaths annually [14], making them the largest contributor to NCD-related mortality. The increasing impact of climate change further exacerbates the burden of NCDs [15]. Heat waves, in particular, increase the risk of heart attacks and strokes among vulnerable populations such as the elderly and those with pre-existing conditions [16,17]. Air pollution is another critical environmental risk factor significantly worsening respiratory diseases like asthma and chronic obstructive pulmonary disease [18]. Wildfires are becoming more frequent and severe due to climate change, releasing harmful smoke and particulates into the air, further compounding the risk for those with existing respiratory conditions [19]. Additionally, extreme weather events such as floods and hurricanes disrupt healthcare access [20], making it harder for individuals with these chronic conditions to manage their symptoms and receive care in real-time. Addressing the impact of climate change on NCDs requires digital technologies and ubiquitous devices such as Internet-connected devices (e.g., smartphones and wearables) which have the potential to transform health systems via rapid response and communication for urgent health crises [21].

NCDs pose a significant health burden in low- and middle-income countries such as India, accounting for 73% of total deaths [22]. Rural Indian populations experience disproportionately higher mortality and morbidity rates due to NCDs [23–26]. Climate change is not only exacerbating NCD risk factors [27,28], but also complicating NCD management among rural communities that face barriers to healthcare access [29,30]. In rural villages around Pune, Maharashtra (Western India), higher frequency of extreme weather events, particularly heat waves [31–33], have increased the risk of heart attacks and strokes, as elevated temperatures create cardiovascular stress [27,29,30,34]. Globally, there is clear evidence that increased temperatures have contributed to higher NCD risk across generations [29,30,35–37] with heat waves worsening already poor air quality across India, leading to respiratory and other chronic diseases [27,34,35]. As a result, it is necessary to develop intersectoral, multi-generational interventions that address the socioecological determinants of health [38–40]. Since many adult-onset NCDs and their determinants originate in childhood and adolescence, it is also imperative to take a life course perspective to minimize intergenerational transmission of NCDs [38,41–43]. To address these challenges, we must integrate climate change adaptation strategies and NCD prevention measures.

In 2022, a report released by the Intergovernmental Panel on Climate Change (IPCC) importantly acknowledged the lack of representation from disadvantaged populations in determining solutions [44]. According to IPCC [44], rural populations in India are at high risk of climate change-related health impacts, particularly due to their reliance on agriculture for livelihood [44–46]. Thus, the use of digital health platforms can play a critical role in amplifying citizen voices and improving health outcomes in disadvantaged communities, where accurate digital epidemiological data are needed to make evidence-based decisions for complex intersectoral problems such as NCDs and climate change. Digital health platforms, including technologies like progressive web applications and digital dashboards, facilitate real-time access to health services and information and provide citizens, decision-makers, and researchers with the tools needed to address critical health challenges [47]. Although there is no silver bullet solution for climate change adaptation and NCD prevention, an opportunity potentially lies in ethically obtaining citizen big data (i.e., ethical surveillance) via citizen-owned digital devices to transform evidence-based decision-making [48].

Ethical surveillance of big data using ubiquitous digital devices (i.e., smartphones) not only engages citizens at all stages of the research process, but also enables citizens to access, pause, or delete their data – an integration of digital epidemiology, digital citizen science, and community-based participatory research, which can be achieved through operationalizing global digital citizen science policy and the Smart Framework [48–54]. For example, digital platforms can enable an early warning system where decision-makers can send alerts for heat waves and floods. This timely information is critical for citizens to adapt to acute events at the household and community level, e.g., carrying water to avoid dehydration, and preparing barriers for flood zones. Moreover, NCD prevention involves strategies to reduce the incidence and impact of chronic diseases by addressing their root causes, e.g., improving awareness of health risks and timely access to relevant information [55].

Inaccurate and invalid data from unreliable sources can result in disjoint and ineffective decision-making [56] – an issue that became apparent in the inadequate response to recent climate disasters across the world, including India [57,58]. Thus, this study aims to adapt, implement, and evaluate a digital health platform for the primary and secondary prevention of NCDs, as well as climate change adaptation in rural India. As described in this protocol, an existing digital health platform [47] will be adapted to connect citizens in rural communities with decision-makers using a progressive web application and a dashboard system to enable near-real-time health services and evidence-based decision-making.

## 2. Methods

### 2.1 Study Design, Participants and Sample Size

This study will utilize a 5-year longitudinal trial with a step-wedge design [59] to adapt, implement, and integrate the Climate Care India digital health platform into an existing 15-year YoUth adolescents’ behaViour, musculoskeletAl heAlth, growth & Nutrition (YUVAAN) cohort study **(Figure 1)** [60].

**Figure 1.**
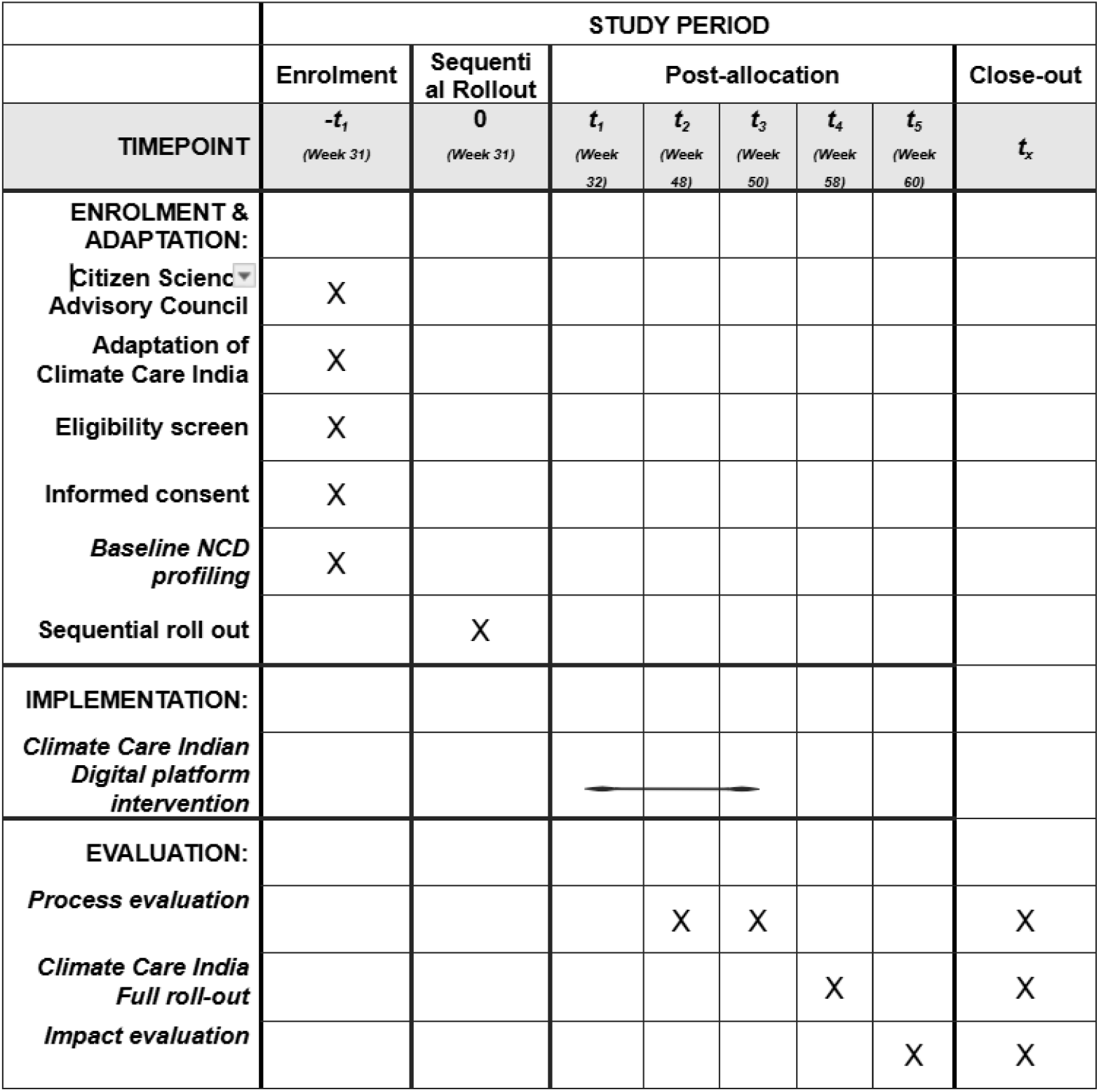
Schedule of Enrolment, Implementation and Evaluation.

The step-wedge design allows the sequential rollout of the intervention across the villages, ensuring that households act as their controls, enhancing external validity [59]. The trial will be conducted across six rural villages participating in YUVAAN that have shown high interest in climate change adaptation (**Figure 2)**.

**Figure 2.**
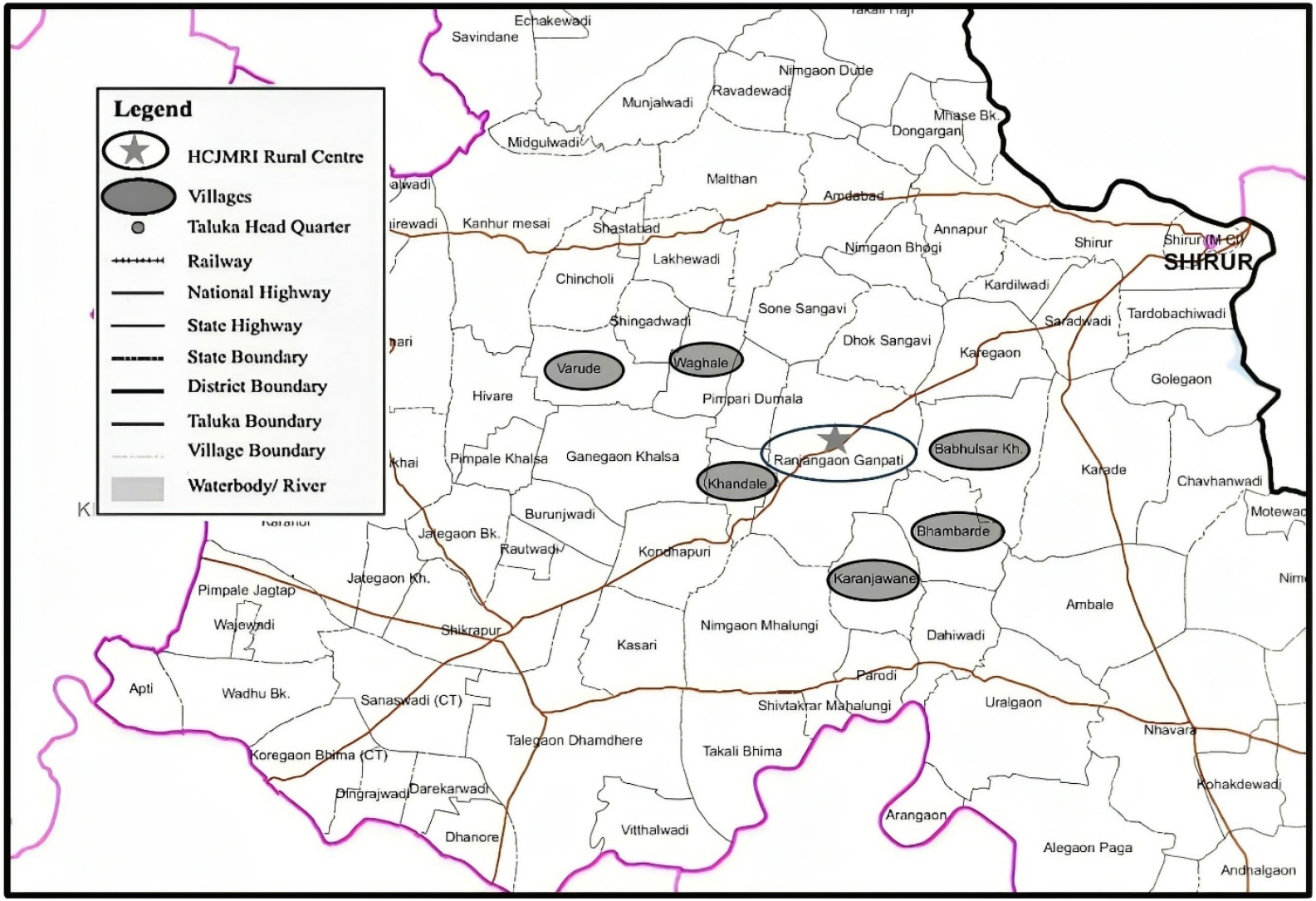
Map of Study Site and Included Villages

The villages are in the Shirur Taluka district, approximately 60 kilometers west of Pune city, Maharashtra, India. These villages include 2,073 households with a total population of 10,398 citizens. A sample size of 978 households (approximately 3,912 citizens) will be recruited, accounting for 37% of the population, based on a calculated sample size to detect a 0.3 effect size with 90% power at a 0.05 significance level. The inclusion criteria for participation require each household to have at least one Internet-connected digital device. Recruitment will focus on households already participating in the YUVAAN study. A convenience sampling technique will be adopted to facilitate ease of participation. Promotional materials, including educational videos in the local languages (Marathi and Hindi), will be used to ensure community awareness. Participation will be confirmed by collecting phone numbers and validating them using ethical global positioning system location estimates.

### 2.2 Climate Care India Digital Health Platform

The existing digital health platform is designed to support rapid decision-making during public health crises [47]. It integrates big data from a progressive web application (PWA) that allows users to assess household COVID-19 risk, request food assistance, and report issues accessing public services. The existing platform features include real-time interactive data visualization, community alert posts, and bidirectional engagement between citizens and decision-makers. This study will adapt the existing platform to manage NCDs and climate change. The new platform, called Climate Care India, will comprise two interconnected components: (i) PWA for citizen engagement: The PWA front-end interface allows a single household member to manage the data of all family members. This approach reduces the burden on both users and healthcare providers. PWA users will be able to create “avatars” to represent household members, which will serve as personalized health accounts. These avatars will store baseline health information and NCD risk factors of each household member. Additionally, the PWA will provide an interface for anonymous communication between healthcare providers and citizens. (ii) digital health dashboard: This will be a back-end digital health dashboard that displays anonymized and aggregated household data for designated decision-makers (i.e., healthcare providers, and community leaders) in easy-to-understand interactive visualization to enable effective public health surveillance and decision-making. Through the back-end dashboard, decision-makers can send community-wide alerts and respond to anonymized household requests or questions in real-time **(Figure 3)**. The platform will be adapted by translating it into local languages, Marathi and Hindi, for use in the rural villages where the study will be conducted.

**Figure 3.**
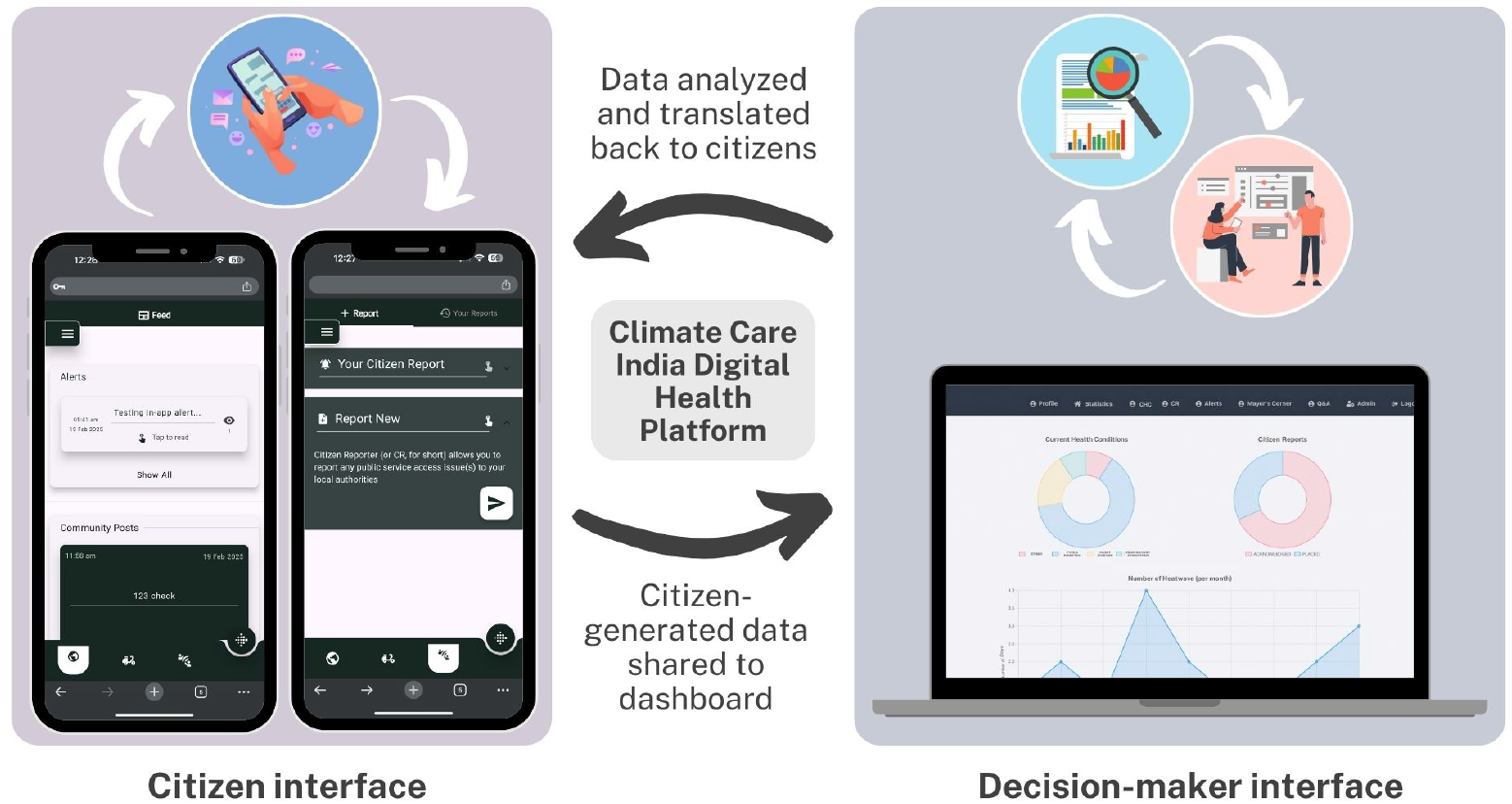
Climate Care India Digital Health Platform

### 2.3 Study Phases

This study will be conducted in three phases, involving six steps to adapt, implement, and evaluate the custom-built Climate Care India platform. **Step 1 (0-6 months)**: A 12-member Citizen Scientists Advisory Council, consisting of two representatives from each village, will be established. The council will receive comprehensive training on the Climate Care India platform and participate in co-design sessions to guide platform adaptation (November 3^rd^, 2025 – August 3^rd^, 2026). **Step 2 (6-30 months):** Adaptation of the platform will occur through an iterative process, including translation into Marathi and Hindi, and rigorously tested with the development team before beta testing with the Advisory Council (August 4^th^, 2026 - August 4^th^, 2028). **Step 3 (30-48 months):** The implementation stage will consist of two steps from months 30 to 48. First, recruitment and baseline NCD risk profiling will be conducted in the six villages from months 30-32. Implementation of the intervention will take place at 8-month intervals between months 32-48, with process evaluations at each interval to assess usability and compliance (August 5^th^, 2028 - February 4^th^, 2030). **Step 4 (48-50 months):** The process evaluations of the platform will be carried out from months 48 to 50 of the study period. Findings from the process evaluations will inform adaptations to the platform (February 5^th^, 2030 - April 4^th^, 2030). **Step 5 (50-58 months):** The full version of the platform will be made available to all households (April 5^th^, 2030 - December 4^th^, 2030). **Step 6 (58-60 months):** An impact evaluation will be conducted to assess the platform’s overall effectiveness in promoting climate change adaptation, improving health behaviours, and reducing NCD risk factors (December 5^th^, 2030 - February 5^th^, 2030).

### 2.4 Data Collection and Analysis

Baseline NCD risk profiles for all participating households will be developed using sociodemographic data including sex, age, household income, highest level of education in the household, employment status and type, and number of people in the household. Health-related variables (i.e., subjective and objective physical health assessments, bone and muscle health assessment, anthropometry), including current NCDs among household members, and behavioural risk factors (i.e., physical activity, screen time, substance use, nutrition, alcohol consumption), will be collected from the YUVAAN cohort study. Data collection for the YUVAAN study commenced on 13 September 2022 and will continue over the next 15 years (i.e., 13^th^ September 2037). Data collection for the Climate Care India study will begin on November 3^rd^, 2025, and end on December 4th, 2030)

Through the Climate Care India platform, household behavioural data will also be collected prospectively to ascertain changes in the NCD risk profile of participating households. These data and profiles will inform personalization and real-time support tailored to individual household members. For example, Climate Care India may track dietary habits, physical activity levels, and environmental exposures to identify emerging health risks. Based on this data, it can provide personalized recommendations, such as nutritional guidance, exercise plans, or targeted health interventions, to support household members in reducing their NCD risk. Household personalization of Climate Care India will be based on machine learning models, large language models, and generative artificial intelligence models [61]. Anonymized data generated from participating households will be aggregated and visualized on the Climate Care India dashboard for real-time clinical decision-making.

Advanced data science techniques will enable health decision-makers to identify evolving community NCD risks and send alerts for adaptation to climate change. External data sources such as climate monitoring data will be integrated with the dashboard to inform decision-makers of severe weather patterns and help guide timely interventions. Traditional statistical analyses will be conducted to evaluate the adaptation of the Climate Care India platform and its impact on healthcare access and outcomes. Agent-based modeling and simulations will assess various scenarios to predict the platform’s future impact on NCD risks and climate adaptation. Additionally, sex and gender-based analyses will be used to evaluate differential impacts across sub-populations at higher risk. All analyses will be conducted using R 4.4.1 statistical software and Python.

### 2.5 Data Management and Ethical Consideration

All data will be securely stored in Amazon relational database cloud servers located in India, adhering to the Government of India’s Digital Data Protection Bill [62]. Data will be encrypted, anonymized, and aggregated, ensuring no personally identifiable information is accessible. Regular data backups will be conducted to protect against data loss. Ethical considerations will include obtaining informed consent from all participating households and implementing stringent data protection measures to ensure household and individual privacy. Ethics approval was obtained for the Adolescent Cohort Study, which has since been renamed to the YUVAAN study. Approval was granted by the Hirabai Cowasji Jehangir Medical Research Institute, India (document attached separately). An ethics amendment will be submitted for the Climate Care India project, which will be embedded into the YUVAAN study framework. Written informed consent was obtained from adults, and written informed assent was obtained from children, with the additional requirement that their parent or guardian also provided written informed consent before the child’s participation.

## 3. Discussion and Implications

This protocol outlines the innovative use of a digital health dashboard for continuous, near-real-time data sharing with decision-makers and healthcare providers. The Climate Care India platform will enable near-real-time surveillance and evidence-based decision-making to address NCDs and climate change adaptation in rural India. This integrated rapid knowledge translation approach [63,64], involving stakeholders such as the research team, knowledge users, and community leaders, aims to apply knowledge throughout the project timeline, enhancing the project’s impact and relevance.

The YUVAAN cohort study will identify prevalent NCD risks in rural India [60], highlighting the urgent need for effective intervention strategies to improve health outcomes [65,66]. In response to these health challenges, this study will adapt and repurpose an existing digital health platform [47] to address NCD risks and empower rural households by enhancing their overall climate change preparedness. This platform will be designed to offer tailored, near-real-time health support, enabling individuals and families in remote areas to manage their health conditions more effectively.

Given the complexity of NCDs, effective management requires dynamic solutions that can be adapted to the evolving nature of individual and contextual (i.e., social, ecological, economic, political) risk factors. Digital health platforms have demonstrated potential in improving NCD management [67,68] by capturing relevant context and enabling citizen/patient-clinician communication to manage existing and emerging risks in real time [69–71]. Digital epidemiology plays a critical role in this process by leveraging real-world data from digital health interventions to identify patterns, predict health risks, and inform intervention strategies [51,72]. In India, digital NCD interventions have been associated with positive health outcomes, including improved medication compliance, communication between patients and clinicians, and reduced disease symptoms [73]. For instance, Kesavadev et al. (2012) conducted a study in Kerala, India involving 1,000 participants with diabetes mellitus [74]. The intervention utilized the Diabetes Tele Management System, which provided telemedicine-based diabetes self-management education and support. The study reported significant improvements in glycemic control and patient satisfaction. Similarly, Ramachandran et al. (2013) conducted a randomized controlled trial in Tamil Nadu and Andhra Pradesh with 537 participants receiving personalized mobile phone messages promoting lifestyle modifications [75]. The study found a significant reduction in the incidence of diabetes in the intervention group compared to the control group, highlighting the potential of mobile-based interventions in diabetes prevention. This evidence underscores the potential of digital interventions in modifying health behaviours and preventing disease progression through digital epidemiological approaches.

The expected outcomes of this project are multifaceted, with the potential for significant impact at the household, community, and health system levels [76]. First, the platform is designed to improve household awareness and understanding of NCDs and related risk factors [77,78]. This will empower families to make informed decisions about their health and behaviour [79]. This aligns with the study’s broader goal of supporting household-level management of NCD risks and climate change adaptation, as well as promoting behaviour modification for improved health outcomes [80].

Climate Care India is also expected to enhance preparedness for climate-related health risks by offering households personalized, real-time prevention and adaptation strategies [81]. By linking citizen-generated big data with personalized interventions [82], the platform will provide citizens with critical health information and decision-making support in real-time [83]. Additionally, the dashboard’s ability to offer anonymized insights to healthcare providers will allow for more informed responses [47], and timely clinical decision-making [47], thereby reducing NCD risks and improving community resilience to adverse climate events [84,85].

Moreover, this project has broader implications for global health services research. Climate Care India’s ethical use of big data for personalized health interventions represents a significant advance in building resilient health systems [86] via digital health technologies, particularly in low-resource settings [87]. Digital epidemiology will play a foundational role in this process by ensuring that real-time data is accurately analyzed and translated into actionable insights [72]. By providing both citizens and decision-makers with epidemiological insights, this platform will contribute to the development of learning health systems – an approach that promotes continuous improvement based on real-world evidence [88]. Furthermore, the project’s potential for scalability across international jurisdictions provides a framework for addressing similar global health challenges in diverse contexts.

## Conclusion

The successful implementation of Climate Care India will not only improve NCD management and climate adaptation at the local level but will also provide a model for future digital health solutions globally. The platform’s ability to deliver near-real-time health support and inform public health decision-making represents a cutting-edge approach to addressing the dual challenges of NCDs and climate change, with the potential for long-term sustainability and scalability. This project will generate valuable insights into the intersection of health and environmental risks, offering a blueprint for precision prevention strategies in the digital age [89].

## Data Availability

No datasets were generated or analysed during the current study. All relevant data from this study will be made available upon study completion.

## Acknowledgments

We acknowledge the entire Digital Epidemiology and Population Health (DEPtH) Lab, Change Research Lab, and the Hirabai Cowasji Jehangir Medical Research Institute teams. The authors also acknowledge the Canada Research Chairs program for their support to the DEPtH Lab.

## Author contributions

Jasmin Bhawra: Conceptualization, Methodology, Investigation, Writing – review & editing. Sheriff Tolulope Ibrahim: Investigation, Writing – Original draft, review & editing. Jamin Patel: Investigation, Writing – review & editing. Anuradha Vaman Khadilkar: Conceptualization. Tarun Reddy Katapally: Conceptualization, Funding acquisition, Methodology, Writing – review & editing.

## Funding sources

This work was supported by the Canada Research Chairs Program

## Notes

### Competing Interest Statement

The authors have declared no competing interest.

### Clinical Trial

NCT05603793

### Clinical Protocols

https://pmc.ncbi.nlm.nih.gov/articles/PMC11812897/

### Funding Statement

Yes

